# Clustering method for spread pattern analysis of corona-virus (COVID-19) infection in Iran

**DOI:** 10.1101/2020.05.22.20109942

**Authors:** Mehdi Azarafza, Mohammad Azarafza, Haluk Akgün

## Abstract

The COVID-19 is outbreak from China and infected more than 131,652 people and caused 7,300 deaths in Iran. Unfortunately, the infection numbers and deaths are still increasing rapidly which has put the world on the catastrophic abyss edge. Application of data mining to perform pattern recognition of infection is mainly used for peparing the spread mapping which considred in this work for spatiotemporal distribution assessment and spread pattern analysis of corona-virus (COVID-19) infection in Iran.

## 1. Introduction

With the spread of the corona-virus (COVID-19) from Wuhan in China [1,2] to all over the world, this virus has become the biggest human challenge. This geographical outbreak of the COVID-19 virus has had a significant impact on the economic, social, and political conditions in various countries which have changed their interactions. Meanwhile, different countries have adopted various restriction procedures (such as closing land, sea and air borders, restricting of international air traffic, etc.) to reduce the infectious effect of COVID-19. However, the COVID-19 outbreak has been very rapid and widespread. Although countries around the world have taken various precautions against the corona disease, it has been rather evident that travel restrictions such as reduction in the trip volume, transit control in the main routes at different levels (e.g., within the towns, intercities, inter-provincial regions and the entire country) is the most important approach to limit this contagious disease [3]. Experiences from different countries have indicated that intercity travelling management can help reduce corona emissions. An understanding of the COVID-19 spreading status in cities can be used for pattern analysis, crisis management, medical services, supply of pharmaceutical-food materials, and specialised forces transportations, etc. Such actions can significantly lead to cost reduction for countries.

February 19, 2020 was the first confirmed corona-virus contamination point in Iran as was reported by the Iranian health authorities which has expanded significantly in the following months [4]. From February 19 to March 22, 2020 (in 33 days of epidemics), the confirmed cases reached up to 21638 persons [5,6] which initiated from Qom and affected the other provinces of Iran [4]. This rapid spread of COVID-19 ranks Iran as the third country with the highest number of confirmed cases after China and Italy up until late March, 2020 [7]. After March 22, 2020, the official COVID-19 confirmed information in provincial level has not been published by the Iran ministry of health and medical education officials and only national information has been reported [6]. However, it is possible to perform a coronavirus spread pattern analysis and contamination risk monitoring based on provincial information which is considered in the study reported herein.

## 2. Materials and Methods

In order to perform a COVID-19 spread pattern recognition in Iran, clustering algorithm [8] and geographical information system GIS [9] were utilised to determine the virus spreading possibility from the starting point (Qom city) to the other parts of Iran. As a cluster analysis, the main task is exploratory data mining to perform pattern recognition of infection development monitoring. As known, clustering is not one specific algorithm, but consists of various algorithms that differ significantly in their understanding of what constitutes a cluster and how to efficiently apply them to solve the task. Clustering can be formulated as a multi-objective optimiser to conduct knowledge discovery of events pattern that is based on an iterative process [8]. As far as GIS mapping [9] is concerned, the main task is to design a visual view based on interactive queries, spatial information analyses and present the results of the operations in the form of a COVID-19 spread pattern map. The partnership of these two approaches enables the utilisation of an effective spatiotemporal distribution tool through provincial-level infection pattern developments in Iran.

## 3. Results and Discussion

Fig. 1 presents the COVID-19 infection situation in Iran in 4 different time periods ranging from February 19 to March 22, 2020, based on cluster data visualisations and GIS mapping based on a k-means algorithm. In Fig. 1, the corona-virus prevalence in Iran for the number 3, 4 and 5 clusters are organised as high-risk, low-risk and moderate risk, respectively, where cluster ‘0’ indicates the low infection risk and cluster ‘5’ indicates high infection risk during February 19 to March 22, 2020. For example in the 3 clustering tables, between February 19 to 26, 2020, the related cluster ‘0’ is low and cluster ‘3’ is high infection risk. Also, in the 5 clustering tables, between February 19 to March 22, 2020, the related cluster ‘0’ is low and cluster ‘5’ is high infection risk in different cities of Iran. Regarding these epidemiological scenarios, it can be inferred that travelling is one of the most effective factors for COVID-19 outbreak in Iran (However, it should be noted that various factors are involved that have caused the complexity of the spread). Fig. 2 presents a Euclidean distance dendrogram after infection clustering and scaling which indicates that there are three widespread infection waves in the country between the cities, and the role of the Tehran city in this prevalence is very significant. Fig. 2 has identified 3 categories for the infection which are shown with as ‘blue’, ‘red’ and ‘green’ colours which shows the cities that might have affected each other after the COVID-19 infection. Fig. 3 illustrates the kernel density estimation (KDE) non-parametric probability density function results that were implemented for the corona disease prevalence in Iran based on spatiotemporal distribution and infection development during the February 19 to March 22, 2020 time period. According to the results of presented by Fig. 3, the Tehran, Qom and Alborz provinces are located within the crisis areas of the country.

**Figure 1.**
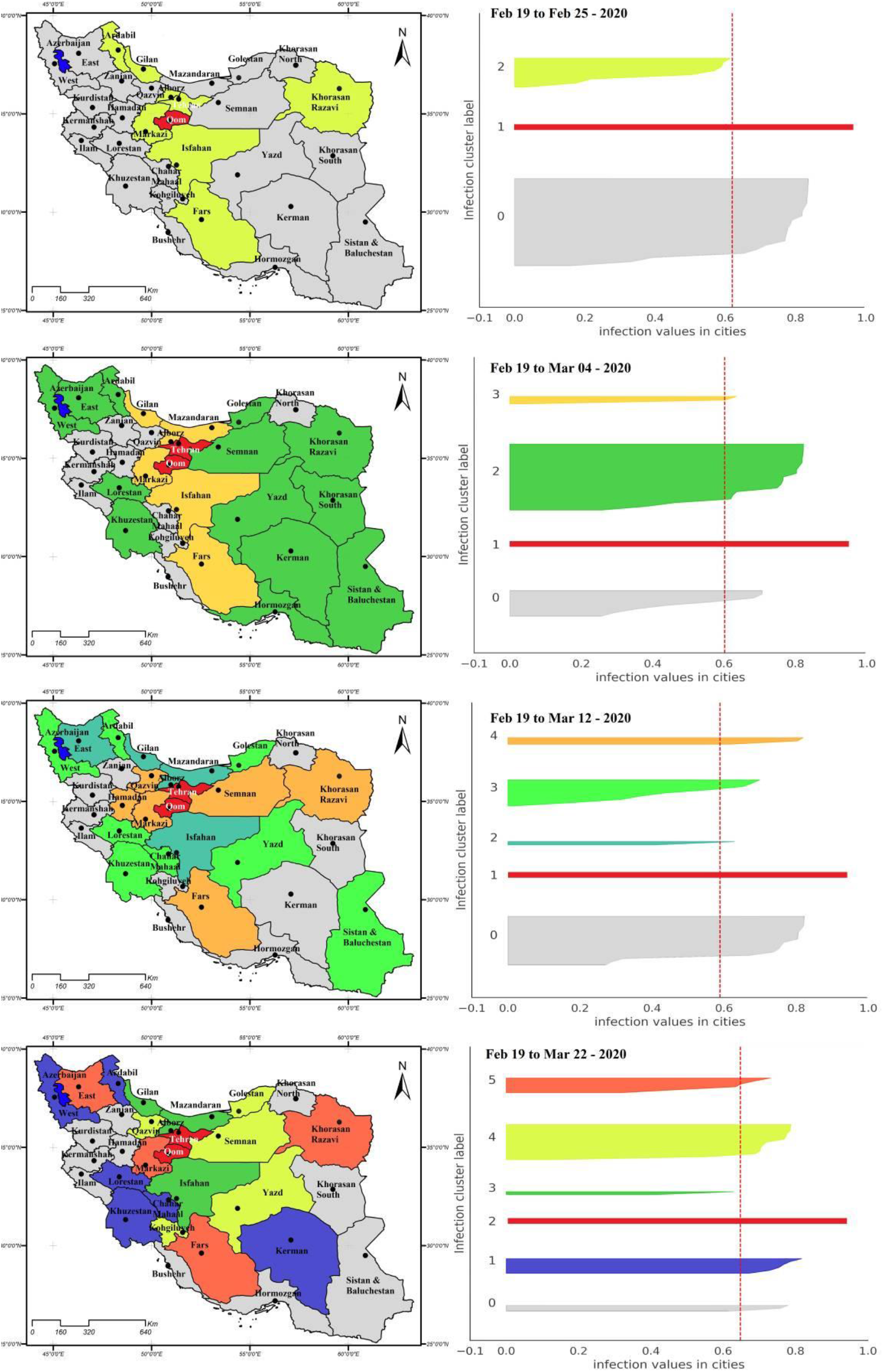
Corona-virus spatiotemporal distribution from February 19 to March 22, 2020 (Data source: [4])

**Figure 2.**
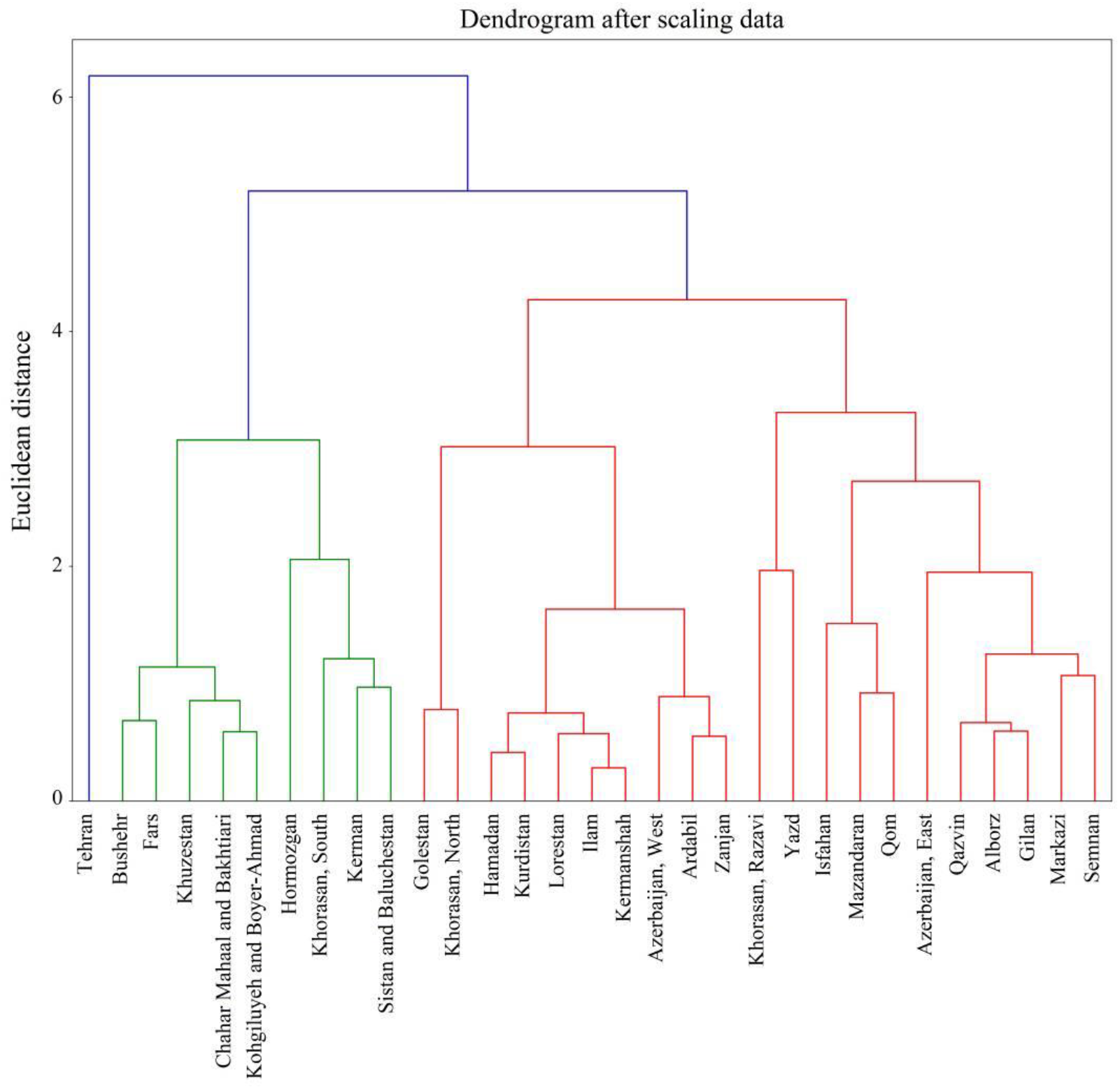
Euclidean distances dendrogram after infection clustering in Iran.

**Figure 3.**
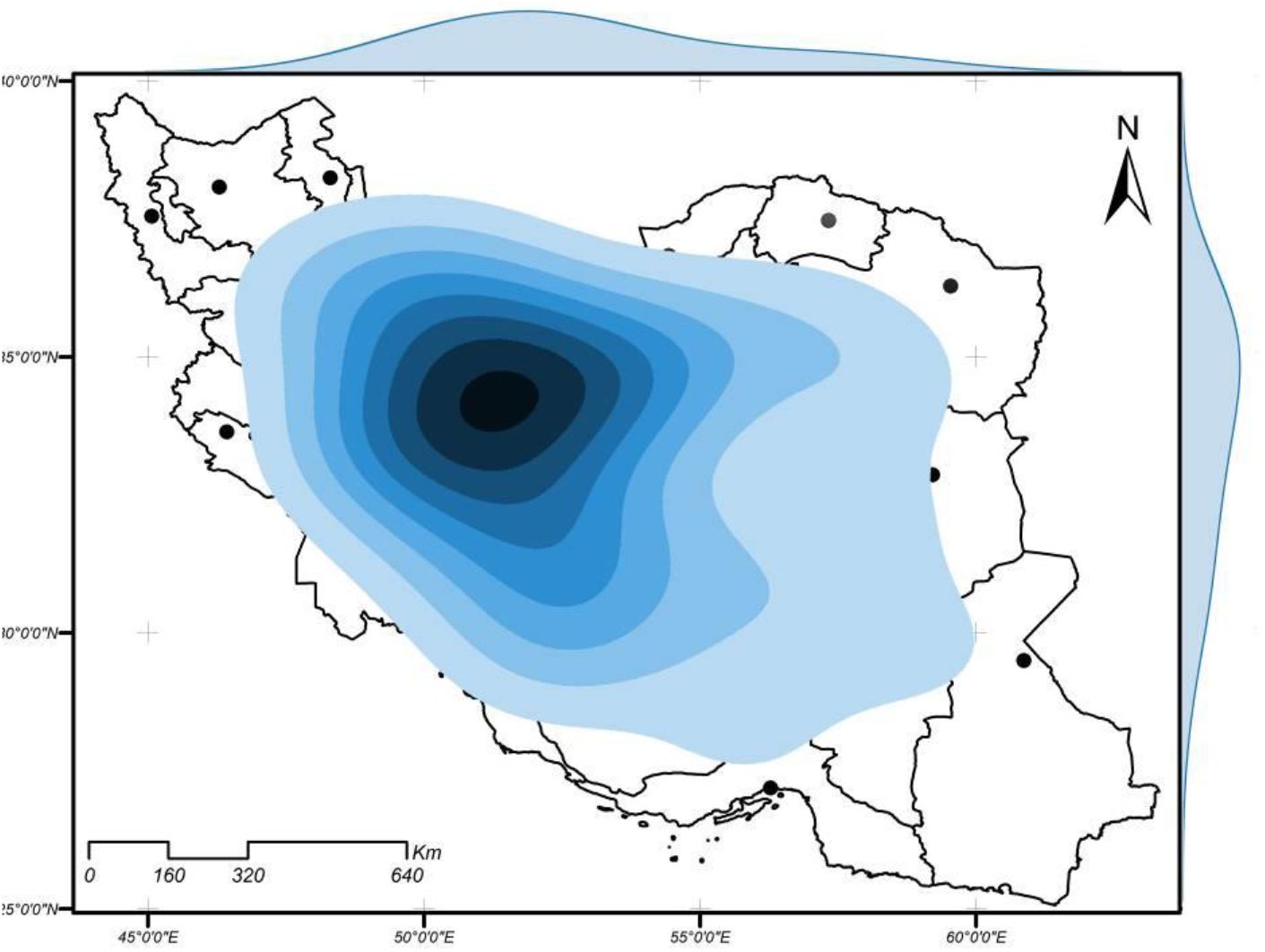
The KDE-based spatiotemporal infection development in Iran (Data source: [4])

After preparing the probability density of the infection maps, the infection pattern with transportation between provinces is estimated by the clustering method. The clustering results that are used to estimate the probabilistic routes of the COVID-19 infection is presented in Fig. 4. The presented figure was drawn by Gephi software. Each ninety identifiers of the desired province, edge and the province connection in terms of the produced cluster is drawn by the k-means algorithm. The relationship of each node with the weights of the province’s cluster labels and the distance proximity to the provinces, the province area and generated clusters are determined by the k-means clustering. In the first week, 11 provinces, in the second week, 27 provinces, and in the third week, all 31 provinces of the country were infected with the transportation system.

**Figure 4.**
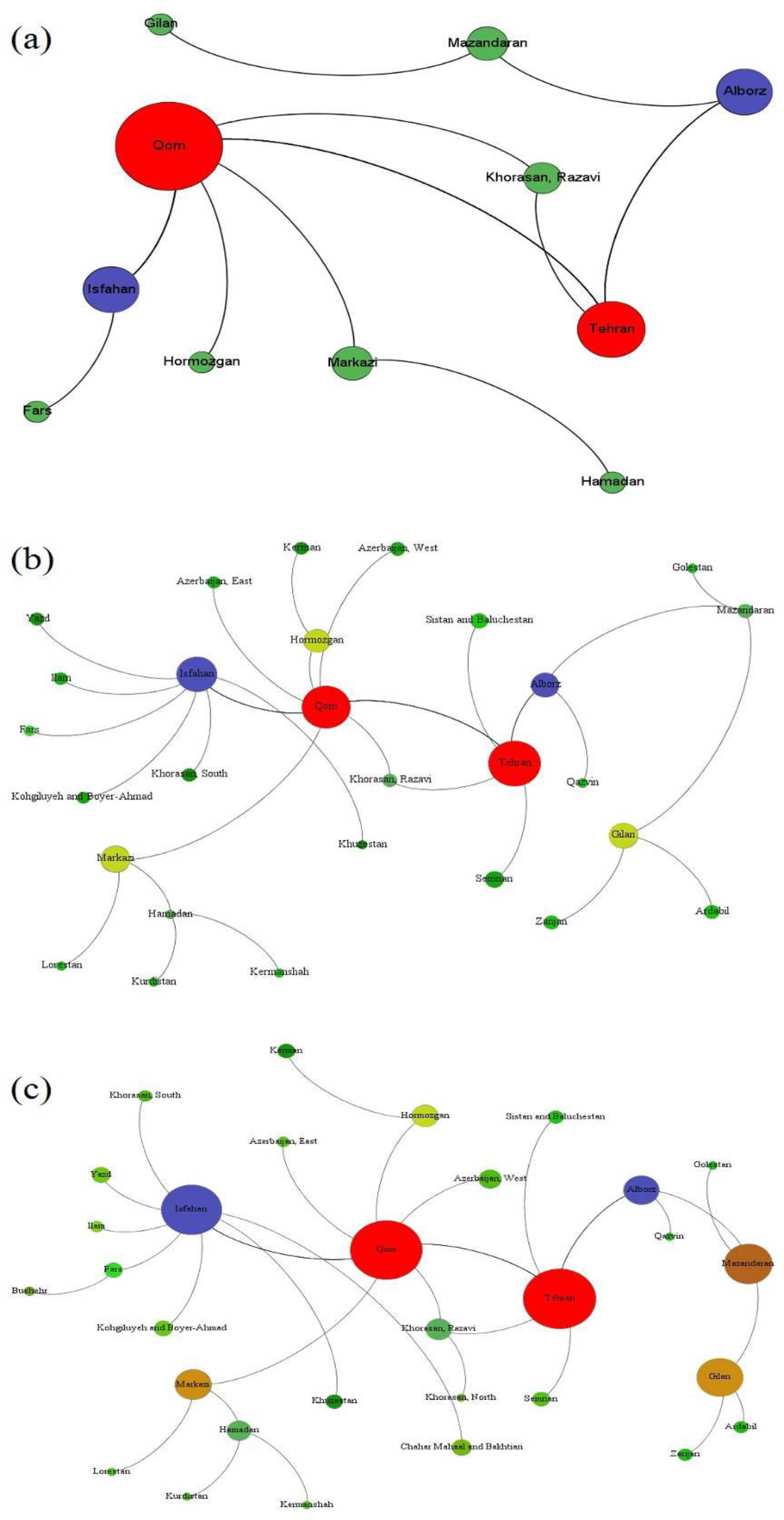
Infectious disease transmission pattern in Iran: (a) February 19 to 25, (b) February 19 to March 4, (c) February 19 to March 22, 2020.

## 4. Conclusion

The presented study attempted to use the clustering analysis method for time series modelling and providing the COVID-19 disease spread pattern in Iran as the provincial level. Provincial data collected from February 19 to March 22, 2020 based on nationally recognised sources (e.g. Iran Ministry of Health and Medical Education; IRNA; ISNA). The data mining to perform pattern recognition of COVID-19 infection in Iran which is mainly applied based on clustering methodology. According to the results Tehran, Qom are the main points for corona-virus expansions. But the Tehran city is the responsible for all infection spread pattern in Iran.

## Data Availability

All data in the paper

## Declaration of Competing Interest

The authors declare that they have no competing interests.

## Funding statement

This research did not receive any specific grant from funding agenciesin the public, commercial, or not-for-profit sectors.

## Notes

### Competing Interest Statement

The authors have declared no competing interest.

### Author Declarations

checked by authors

